# CONSILIENCE-GWAS: A Web Resource for Parsed Heritability & Polygenic Score Analysis of Human GWAS Using Heterogeneous Functional Genomics Data

**DOI:** 10.1101/2025.10.24.25338727

**Authors:** Rameez Syed, Chelsie E. Benca-Bachman, Spencer B. Huggett, John E McGeary, Jason A. Bubier, Heidi Fisher, Alex Berger, Erich Baker, Elissa J. Chesler, Penelope A Lind, Rohan H C Palmer

**Affiliations:** Behavioral Genetics of Addiction Laboratory, Emory University, Atlanta, GA, USA; Psychiatry & Human Behavior, Brown University, Providence, Rhode Island, USA; Providence VA Medical Center, Providence, Rhode Island, USA; The Jackson Laboratory, Bar Harbor, Maine, USA; Belmont University, Nashville, Tennessee, USA; Psychiatric Genetics, QIMR Berghofer, Brisbane, QLD, Australia; Department of Psychology, Emory University, Atlanta, GA, USA

**Keywords:** GWAS, Functional genomics, Cross-species, LDSC, Enrichment analysis

## Abstract

**Motivation:** Addiction-related GWASs are increasingly important in understanding risk for substance use and disorders (SUDs). GWASs have begun to identify loci associated with SUDs and other complex traits, but most associated loci necessitate functional evidence to translate and prioritize. Data depots of functional evidence in tissues, cell types, and experimental paradigms across organisms hold significant promise when integrated with GWAS. Model organisms can provide crucial evidence for gene roles in behaviors; however, public facing resources connecting these data types and allowing for a comprehensive assessment and integration of functional evidence are limited. We address this technological gap by introducing the CONSILIENCE-GWAS analytical webtool that simplifies the aggregation, analysis, and visualization of the enrichment of model organism functional data in human GWAS.

**Results:** We developed CONSILIENCE-GWAS to investigate how GWAS results are enriched by functional evidence within and across model systems. CONSILIENCE-GWAS incorporates enrichment pipelines for LD Score Regression and gene-set based polygenic score creation using user-supplied GWAS results and genotype data. CONSILIENCE-GWAS integrates with GENEWEAVER-GWAS via APIs, allowing end-users to upload, curate, compare, analyze, and share their results. GENEWEAVER integration facilitates the efficient identification of enriched gene sets across species and improves the interpretation of non-coding variants in tissue-specific contexts. We demonstrate features of CONSILIENCE-GWAS using publicly available data from a published enrichment analysis of alcohol consumption in *Mus musculus* translated to human GWAS of alcohol use and disorder.

**Availability and Implementation:** CONSILIENCE-GWAS is freely available on the web at https://consilience.bgalab.emory.edu.

## 1. Introduction

Cross-species genomic analysis has become a primary focus in studying the genetic basis of complex human traits, including substance use disorders (SUDs) (Brasher, et al., 2023). By using the knowledge of genetic conservation, researchers have identified risk genes and pathways in model organisms, like rodents, as their genetic and behavioral traits are often similar to those of humans (Huggett, et al., 2021). For example, the genes and pathways involved in alcohol-related behaviors in mice (such as consumption, preference, withdrawal, neurotransmitter signaling, stress response, and reward), are also linked to the alcohol use disorder (AUD) in humans (Agrawal, et al., 2012). This approach helps prioritize and interpret genome-wide (i.e., p<5e-8 or q-value < 0.05) and nominally (p<1e-4 or q<0.10) significant genes and genetic loci for further research, which could lead to new treatment strategies for AUD.

Model organisms, especially rodents and primates, offer a powerful system for functionally interrogating conserved genetic pathways (Palmer, et al., 2021). For instance, genes regulating alcohol consumption, withdrawal, and reward behaviors in mice have clear parallels in human addiction neurobiology (Huggett, et al., 2024). The shared genetic mechanisms of alcohol-related behaviors in animals and humans emphasizes the translational value of the animal models in studying these complex traits (Benca-Bachman, et al., 2023). By combining animal model data with human genetic findings using tools like LD Score Regression (LDSC) (Bulik-Sullivan, et al., 2015), researchers can partition trait heritability into functional categories. Genomic partitioning aids in the identification of tissue and cell-specific enrichment patterns (Finucane, et al., 2015), making it easier to understand the genetic architecture of SUDs and other complex traits. This framework effectively links genetic variations to their biological functions, allowing researchers to prioritize key genes and pathways for future exploration (Finucane, et al., 2015). However, the process of gathering, harmonizing and exposing model organism data to human GWAS analysis is cumbersome and a framework systematically integrating this cross-species evidence with human GWAS data is needed to make this operation accessible to researchers. Current frameworks that harmonize GWAS summary data, and functional evidence within and across species, focus on evaluating the relative contribution of GWAS effects in sets of genes, biological pathways, cell types, or loosely defined groups of genetic loci (Palmer, et al., 2021). Several tools (e.g., methods, software/tool, databases) exist that facilitate multiDomics and crossDspecies research (e.g., functional mapping and annotation of genetic associations (FUMA) (Watanabe, et al., 2017), single-cell-to-GWAS (Li, et al., 2025), GWAS-Singe Cell (Li, et al., 2025), Brain eQTL Almanac (BRAINEAC) (Ramasamy, et al., 2014) GeneWeaver (Li, et al., 2025), and dbGAP (Mailman, et al., 2007; Tryka, et al., 2014)). However, these tools exist as separate resources, which limits hypothesis generation. Consequently, placing these data in the context of GWAS analysis remained somewhat limited.

Recognizing the significance of exploring shared genetic mechanisms between animal models and humans in addiction research, we introduce CONSILIENCE-GWAS. This web-based tool is designed to automate the complex process of partitioning heritability across various tissue and cell-type genomic annotations, leveraging the LDSC analysis pipeline. CONSILIENCE-GWAS runs on a Linux server equipped with up-to-date versions of Ubuntu, Apache (2.4.52), and PHP (8.1.2). The website follows the Model-View-Controller (MVC) design framework. The Model handles major computational functions on the backend—i.e., generating output result files from input data— implemented in R and Python using tools such as LDSC, Bioconductor, and various visualization packages. The frontend interface is built with HTML, CSS, and JavaScript, while PHP supports data display and execution of R and Python scripts.

Users of CONSILIENCE-GWAS can upload genes from gene sets or select GeneWeaver (Baker, et al., 2016) gene set IDs and summary-level GWAS data for their trait(s) of interest. At this time, analyses are restricted to GWAS summary data from individuals of European ancestry. CONSILIENCE-GWAS can also estimate the enrichment of functional and experimental data using genotype-phenotype summary data from the Mouse Phenome Database [RRID:SCR_003212; (Bogue, et al., 2023; Bogue, et al., 2023)]. It also allows users to incorporate functional annotations from resources such as the Genotype-Tissue Expression project (GTEx) (Consortium, et al., 2015) and the Roadmap Epigenomics Consortium (Finucane, et al., 2018). This platform enables researchers to identify tissues and cell types that contribute significantly to the trait heritability. It combines insights from cross-species gene sets and human-specific epigenetic data, providing a powerful framework to understand the genetic basis of complex diseases and their underlying biological mechanisms to the degree that the psychology and underlying biology of the species and traits overlap (Figure 2). In the proceeding sections we describe how to employ CONSILIENCE-GWAS using publicly available and privately owned data. We also showcase its gene set development tools from GeneWeaver, which can be used to prioritize genes and loci for analyses.

**Figure 1.**
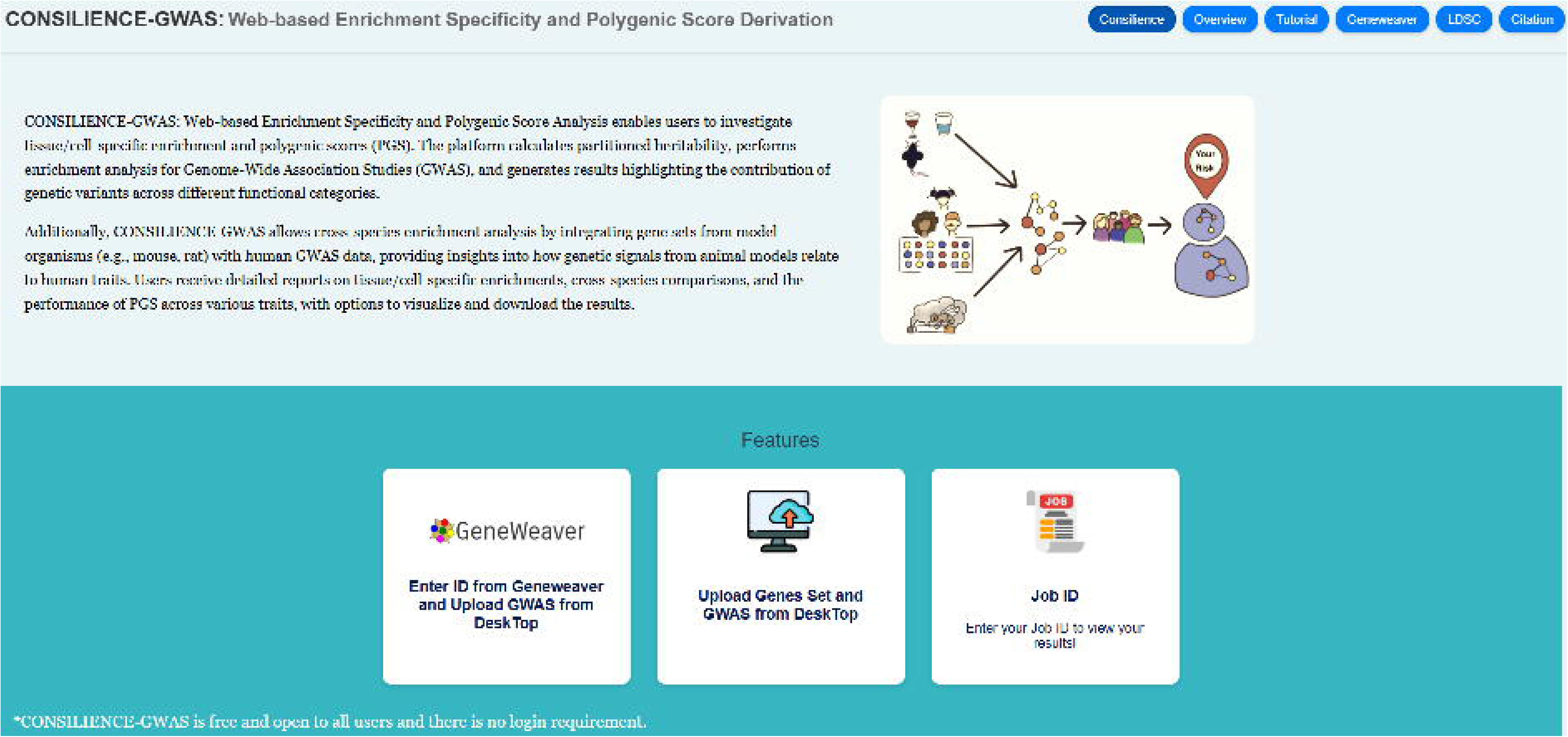
Overview of the CONSILIENCE-GWAS Web Server Architecture. CONSILIENCE-GWAS is a web-based platform that enables users to perform tissue- and cell-specific heritability enrichment, cross-species genomic integration, and polygenic score analyses through an automated and interactive workflow.

**Figure 2.**
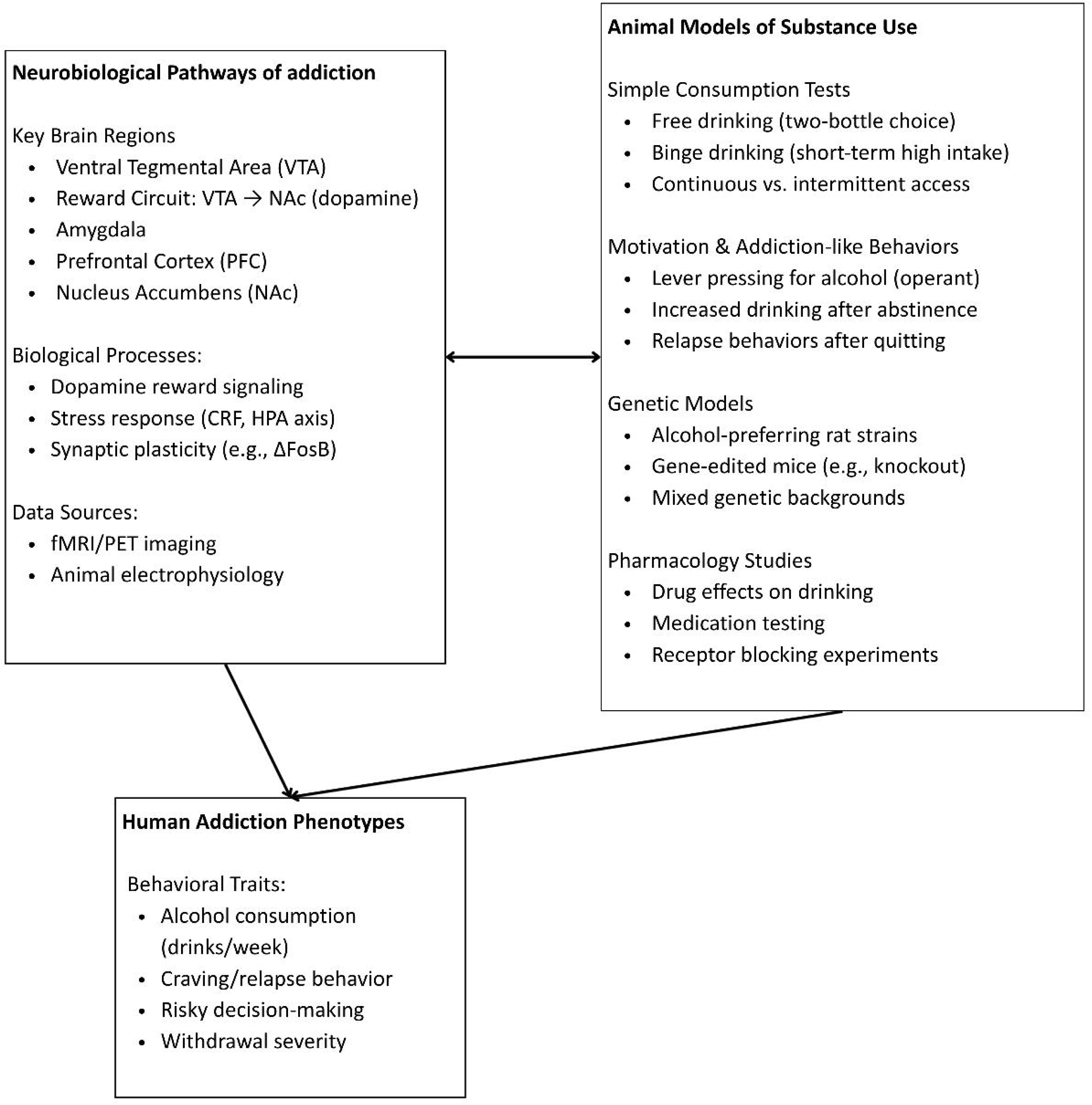
Example Cross-Species Integration of Evidence using Substance Use Disorder Genetics. Neurobiological mechanisms in human (left) and animal models of substance use related behaviors (right) inform the interpretation of human GWAS findings (bottom). Overlapping pathways (arrows) highlight conserved biology across species

## 2. Methods

### Overview of the CONSILIENCE-GWAS Pipeline

The CONSILIENCE-GWAS webtool is a user-friendly interface that provides end-users access to a sophisticated workflow that facilitates the use of several cutting-edge tools to (1) determine enrichment in GWAS summary statistics using gene sets, (2) the derivation of single nucleotide variant weights for creating polygenic scores, and (3) annotation-informed enrichment analyses using cell/tissue data. The pipeline (see Figure 3) is executed on a dedicated high performance computing cluster that allows submitted jobs to be run in parallel using multiple cores. CONSILIENCE-GWAS leverages two key analytical tools: PRS-CS, which estimates single nucleotide polymorphism (SNP) effect sizes and generates polygenic scores based on GWAS summary statistics (Ge, et al., 2019), and LDSC, which is used to estimate tissue-level enrichment of trait heritability by testing whether genes derived from model organisms are disproportionately located within functionally annotated genomic regions (Bulik-Sullivan, et al., 2015). CONSILIENCE-GWAS also supports cell-type-specific enrichment by incorporating reference annotations from public resources such as GTEx (Consortium, et al., 2015) and the Franke lab (Finucane, et al., 2018), enabling users to identify the most relevant cell types contributing to trait heritability. As described in detail below, the analysis process involves three straightforward steps: (1) uploading GWAS summary statistics, (2) uploading a manually curated gene set or an available public or shared gene set from GeneWeaver, and (3) visualizing the results of the heritability enrichment analysis. Users do not need login credentials to access the tool. Users can also download their results as comma-separated value files with an additional option to visualize significant genes as an automatically generated Manhattan plot. A unique job ID is provided to allow users to retrieve their outputs. A complete tutorial for CONSILIENCE-GWAS is available on our website (https://consilience.bgalab.emory.edu) under the Tutorial section. This paper describes a general use case.

**Figure 3.**
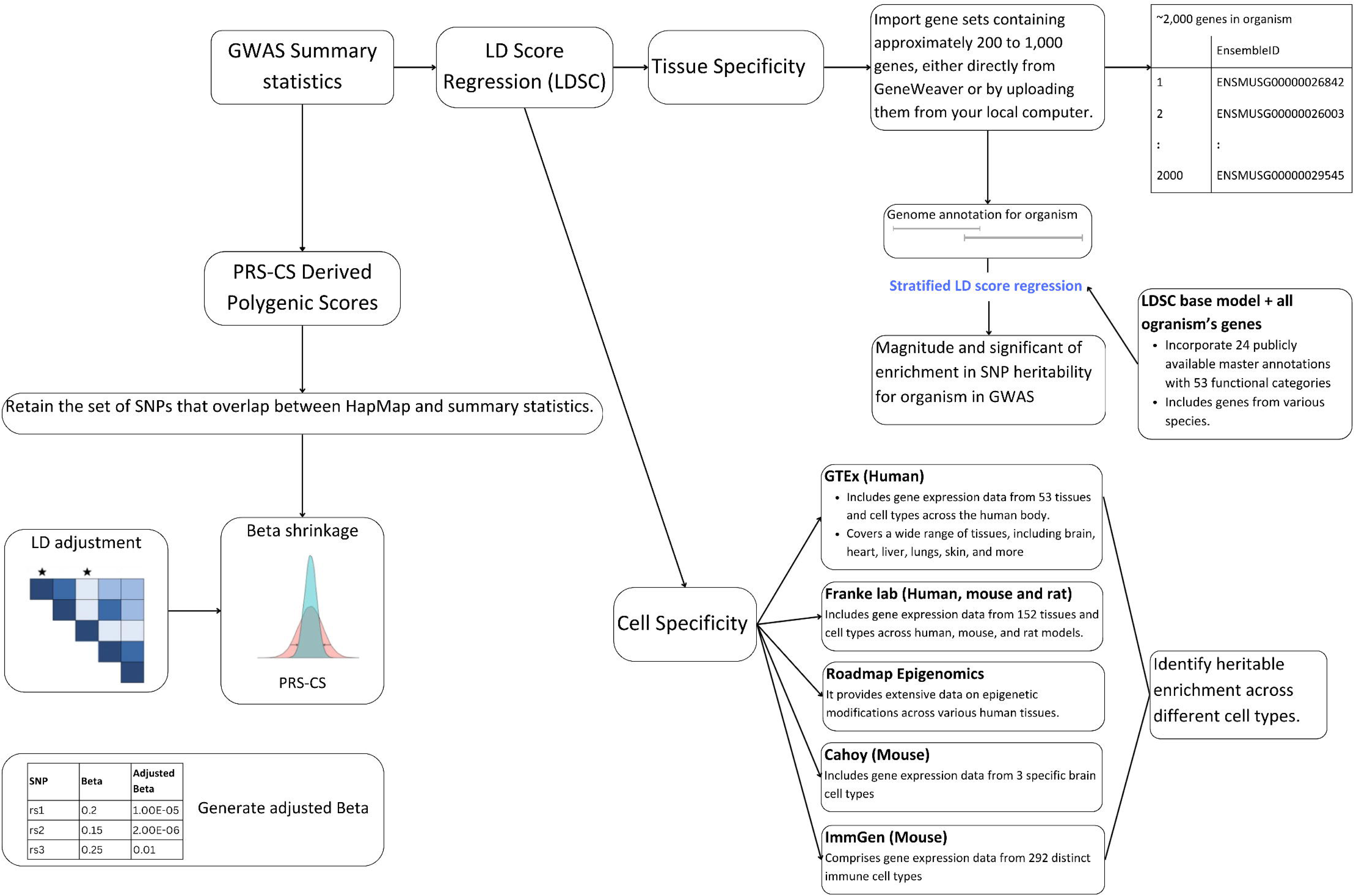
Scope and Features of CONSILIENCE-GWAS. The CONSILIENCE-GWAS server offers three main features: (i) PRS-CS, which allows users to upload GWAS summary statistics and estimate adjusted beta values overlapping with SNPs in organism-specific gene sets; (ii) LD Score Regression (LDSC) for Tissue Specificity, enabling users to upload organism-specific gene sets or select GeneWeaver type IDs to estimate tissue-specific enrichment with human GWAS summary statistics; and (iii) LDSC for Cell Specificity, allowing users to assess cell-specific enrichment using public organism datasets, such as GTEx and the Franke lab resources, in combination with human GWAS summary statistics, without the need to upload organism data.

#### 2.1 Input Data

CONSILIENCE-GWAS requires at least two integral pieces of information: (1) Function evidence in the form of ‘A Priori Gene-sets’ and (2) the full complement of GWAS summary statistics using array, sequencing, or imputed variant data. The pipeline is designed to accept both pieces of information in predefined formats that are typically provided as output from public databases (e.g., Geneweaver (Baker, et al., 2012), Expression Atlas (Papatheodorou, et al., 2020), etc.) and GWAS analysis tools e.g., PLINK (Purcell, et al., 2007) and GCTA (Yang, et al., 2011). A description of the components and features are as follows:

##### 2.1.1 Gene set Input for CONSILIENCE-GWAS Analysis

To perform enrichment analyses, users must compile gene sets relevant to their trait of interest. The platform accepts two input formats: (1) GeneWeaver-generated gene set IDs or (2) manually curated gene lists in standard file formats (.txt). We strongly recommend utilizing GeneWeaver ((Baker, et al., 2012); https://www.geneweaver.org) due to its comprehensive collection of curated gene sets from nine supported species, along with annotation, and analysis tools (Figure 4). As shown in Figure 4a, users can search GeneWeaver for relevant studies, such as "binge drinking", and compare/contrast identified gene sets from different studies to create larger sets (i.e., through unions) or more targeted sets (i.e., through intersection operations) (see Figure 4b). While CONSILIENCE-GWAS was designed to enable cross-species integration, it is equally effective for analyses within a single species; GeneWeaver hosts a vast amount of human genomic data, making it an ideal resource for human-focused studies as well. CONSILIENCE-GWAS processes submitted gene sets using biomaRt (Durinck, et al., 2005) to map genes to their human orthologs, ensuring consistent genomic alignment with GWAS summary statistics. This enables cross-species integration for organisms such as mice and rats. Users can consult the GeneWeaver tutorial or original publication (Baker, et al., 2012) for step-by-step guidance on building, curating, and formatting gene sets compatible with CONSILIENCE-GWAS.

**Figure 4.**
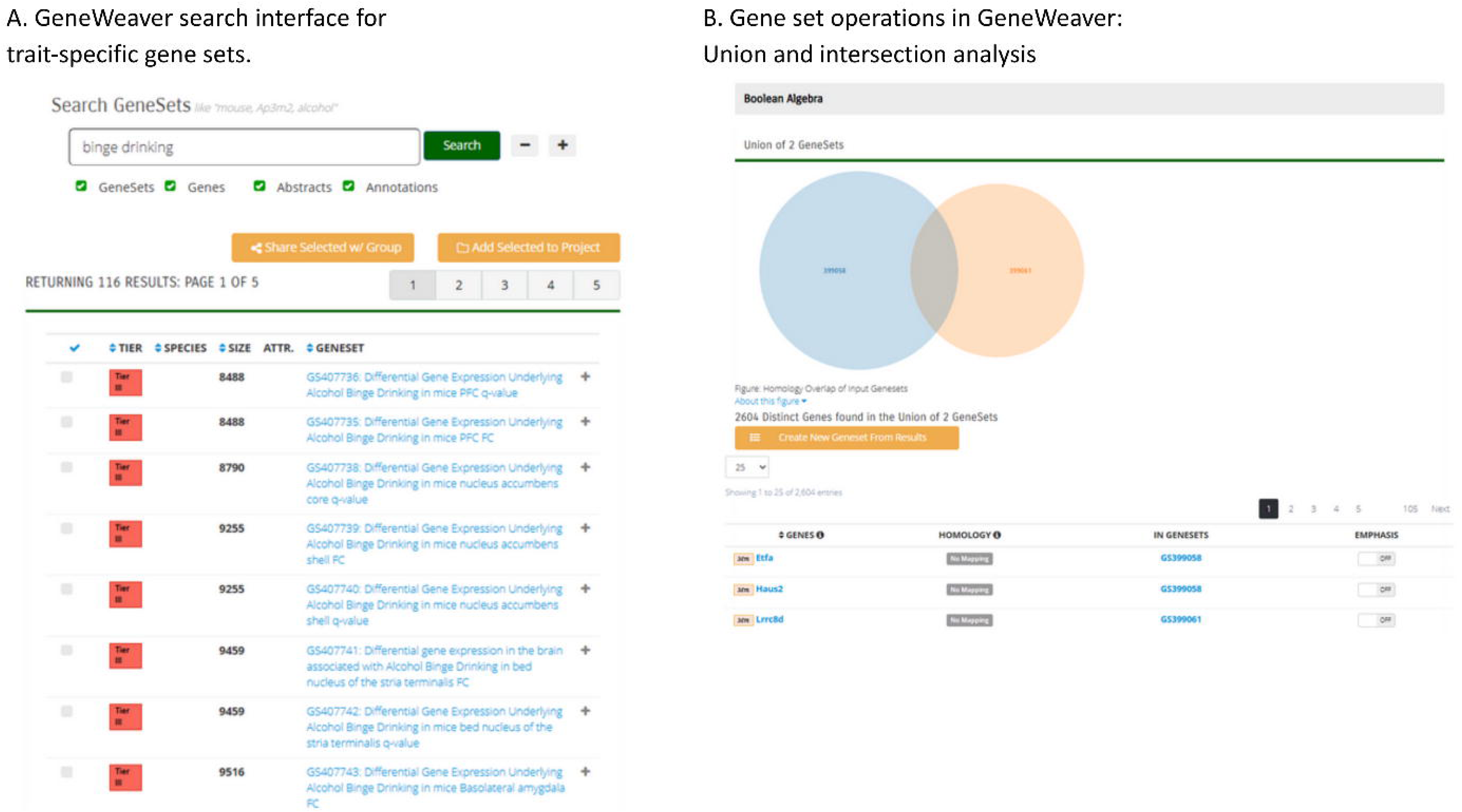
Gene Set Construction Using GeneWeaver for Enrichment Analyses in CONSILIENCE-GWAS. The process of building trait-relevant gene sets using GeneWeaver. (a) Users can search for studies related to specific traits (e.g., "binge drinking") and explore associated gene sets. (b) GeneWeaver allows users to merge gene sets (union) or find overlapping genes (intersection) to refine analyses. These curated gene sets are then submitted to CONSILIENCE-GWAS for enrichment testing against GWAS data.

##### 2.1.2 Human GWAS Summary Statistics Identification and Quality Control

Once a user has selected their gene set(s) of interest, the next step is identifying their target GWAS for analysis. CONSILIENCE-GWAS includes an interface for users to upload and analyze their GWAS summary statistics file. This file must contain columns for SNP rsID, CHR, POS, effect allele, non-effect allele, beta, SE and P-values. Users are also required to provide the sample size for the GWAS file. To ensure the GWAS datasets comply with the LDSC software input requirements (Bulik-Sullivan, et al., 2015), CONSILIENCE-GWAS performs automated quality control procedures on the provided GWAS data. First, CONSILIENCE-GWAS will filter out SNPs with ambiguous allele information, such as A/T or G/C, which can be prone to strand errors (Anderson, et al., 2010). Second, CONSILIENCE-GWAS will restrict the set of variants in the GWAS file to include only HapMap variants, to reduce errors from poor data quality (2005).

#### 2.2 CONSILIENCE-GWAS analysis models

CONSILIENCE-GWAS utilizes two well-established software packages, LDSC v1.0.1 (Bulik-Sullivan, et al., 2015) and PRS-CS (Ge, et al., 2019), that carry out two distinct analyses: assessing cell/tissue specificity enrichment and deriving polygenic scores, respectively. Users are advised to review LDSC and PRS-CS to comprehend the workflow of each of these methods. LDSC is used to assess tissue and cell-type-specific enrichment by estimating whether trait-associated genetic variants are concentrated within annotated genomic regions. This is achieved through Stratified LD Score Regression (S-LDSC version 1.0.1), an extension of LDSC that models the contribution of specific annotations (e.g., enhancer elements or brain tissue chromatin marks) to SNP-based heritability (Finucane, et al., 2018). On the other hand, PRS-CS, derives PGS by using summary statistics from a GWAS to estimate posterior effect sizes through a Bayesian framework. This method accounts for the linkage disequilibrium (LD) structure among SNPs, adjusting effect sizes to generate more accurate polygenic risk scores (Ge, et al., 2019). By incorporating both tools, CONSILIENCE makes it easier for researchers not familiar with programming to ask novel questions about enrichment within and across species for related or unrelated behaviors and traits, and to examine polygenic scores.

Building upon the foundational methodologies described, CONSILIENCE-GWAS further extends its analytical capabilities by conducting three types of joint model analyses. Utilizing LDSC, the tool combines multiple molecular annotations from GeneWeaver gene sets to evaluate their contributions to the heritability of diseases or traits, as detailed below:

**First model:**

This model allows for the estimation of heritability attributable to both unique and overlapping components of the provided gene sets.

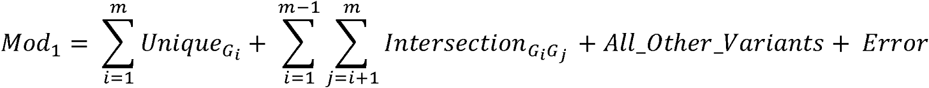

**Summation for Unique Terms:**

⍰ The term 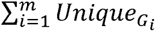 iterates through all *G_i_* groups in the sequence from 1 to m.

**Summation for Intersection Terms:**

⍰ The nested summation 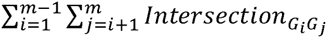 captures all possible pairwise intersections between groups *G_i_* and *G_j_*, where j > i.

**Second model:**

This model assesses the joint contribution of all combined gene sets provided (or from Geneweaver) along with other variants.

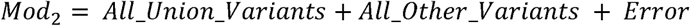

**Third model:**

This model evaluates the joint contribution of each individual gene set provided (or from Geneweaver) in conjunction with other variants.

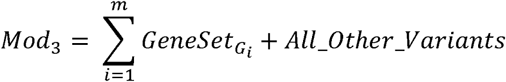

##### 2.2.1 How to Perform Partitioned Heritability Analyses Using CONSILIENCE-GWAS

The primary usage of CONSILIENCE-GWAS is for partitioned heritability analysis using S-LDSC. To illustrate how this works, we applied S-LDSC via CONSILIENCE-GWAS to human GWAS summary statistics to quantify the enrichment of SNP heritability across gene sets derived from GeneWeaver. CONSILIENCE-GWAS automatically deploys S-LDSC’s basic model which adjusts for 53 functional categories, such as conserved elements, enhancers, and genomic architecture (Finucane, et al., 2015).

Users should note that the number of uploaded gene sets determines the type of partitioned heritability models that CONSILIENCE-GWAS will execute. If only a single gene set is provided, the platform performs a standard enrichment analysis estimating the proportion of SNP heritability attributable to that specific annotation, controlling for the 53 baseline categories. In this case, no union or intersection models are run, and only Model 3 (individual gene set) is applied. However, when users upload two or more gene sets, CONSILIENCE-GWAS automatically enables all three modeling strategies. This includes Model 1, which partitions heritability into unique gene set components, pairwise intersections, and remaining SNPs; Model 2, which evaluates the combined enrichment of all uploaded sets; and Model 3, which tests each gene set individually. These joint modeling approaches allow users to explore additive, overlapping, or distinct contributions of gene sets to trait heritability. It is important to note that the runtime averages ∼5.5 hours but may extend up to ∼18 hours for larger datasets, as the computational complexity of the analysis scales with the number of uploaded gene sets; we strongly advise limiting inputs to five gene sets per analysis to ensure efficient processing.

Annotation and LD score files are generated using the make_annot.py and ldsc.py scripts provided in the LDSC software package (Bulik-Sullivan, et al., 2015), following standard parameters. The analysis will incorporate stringent quality control measures, including restriction to HapMap3 SNPs (2005) with imputation INFO scores ≥ 0.9 and minor allele frequency (MAF) > 5%. Users can specify whether LD scores are computed using populations of interest, such as European ancestry samples from the 1000 Genomes Project Phase 3 (Consortium, 2015), with SNP weights and reference data obtained from https://github.com/bulik/ldsc. Currently CONSILIENCE-GWAS can only handle European ancestry sample; future versions will include additional reference samples. Statistical significance for partitioned heritability analyses will be evaluated using a false discovery rate (FDR) threshold of q< 0.05, and enrichment will be quantified as the ratio of the proportion of SNP heritability attributed to an annotation to the proportion of SNPs in that annotation.

##### 2.2.2 Cell- and Tissue-Type-Specific Analysis

In addition to analyses using user-provided gene-sets, CONSILIENCE-GWAS also execute cell-specific heritability enrichment analyses using LDSC-SEG (Finucane, et al., 2018). LDSC-SEG evaluates whether genes with high expression specificity in particular cell types or tissues contribute disproportionately to SNP heritability in the provided GWAS summary data. Specifically, CONSILIENCE-GWAS uses precomputed LD score annotation files (Finucane, et al., 2018), which include 205 tissue and cell-type annotations curated by the Franke laboratory (Pers, et al., 2015). These annotations reflect diverse human biological systems including the brain, immune system, endocrine organs, and musculoskeletal tissues. The included annotations were selected based on robust expression specificity and biological relevance across common complex traits. In addition, the tool integrates 396 chromatin state annotations from the Roadmap Epigenomics (Kundaje, et al.) and ENCODE Projects (Consortium, 2012), which capture tissue-specific regulatory activity such as histone modifications and open chromatin regions. These annotations provide complementary information to expression-based models by capturing the regulatory potential of non-coding genomic regions. To extend translational relevance to neurogenetic studies, CONSILIENCE-GWAS also includes annotations for three major mouse brain cell types—astrocytes, neurons, and oligodendrocytes—based on transcriptomic profiles (Cahoy, et al., 2008), chosen for their well-characterized roles in CNS function and potential overlap with human brain tissue architecture.

We also analyzed 10 aggregated cell-type groups (e.g., adrenal/pancreas, central nervous system, cardiovascular, immune/hematopoietic, liver, and skeletal muscle) to evaluate enrichment of SNP heritability across broader biological systems (Finucane, et al., 2018). Bonferroni correction was applied to determine statistical significance for these 10 categories (P < 0.05). These analyses will help the users to identify cell types and tissues with enriched genetic signals for the traits under study.

#### 2.3 Using CONSILIENCE-GWAS to Derive Polygenic Scores

CONSILIENCE-GWAS leverages PRS-CS (Ge, et al., 2019) to estimate posterior effect sizes (SNP weights) for variants located within user-provided gene sets for participants of European ancestry. PRS-CS is a computationally advanced tool that provides a robust framework for PGS calculation. Compared to traditional clumping and thresholding methods (Purcell, et al., 2007), PRS-CS provides improved predictive accuracy, especially in large-scale datasets, because it explicitly models the LD structure among SNPs and incorporates assumptions about genetic architecture. In genetic studies, PRS-CS has been shown to outperform many other PGS derivation methods by leveraging both GWAS summary statistics and LD correlation data specific to ancestral populations (Ge, et al., 2019; Privé, et al., 2020). CONSILIENCE-GWAS deploys PRS-CS-auto to estimate posterior effect sizes while automatically tuning hyperparameters (Ge, et al., 2019).

Given evidence that a GWAS is enriched for a gene set(s), users can opt to interpret and use CONSILIENCE-GWAS’s derived SNP-weights to create gene set-specific PGSs. Polygenic score derivation is an automated feature that will run for each analyses requested. Users are provided with an updated GWAS summary statistics file that includes weights for each model parameter that is tested in the partitioned heritability analysis. Users can then take scores to use in an independent dataset to construct a PGS using the PLINK (Purcell, et al., 2007) --score option. Average runtime observed for deriving PGS weights is 3.5 days +/- 12 hours.

## 3. Result

### Example CONSILIENCE-GWAS Use Case: Enrichment of Alcohol Addiction Loci in Cross-Species Gene sets

To illustrate the application of CONSILIENCE-GWAS, we used GWAS summary statistics for Drinks per Week (DPW) from the GSCAN study (European ancestry; N = 537,349, excluding 23andMe samples) (Liu, et al., 2019), whose analysis identified multiple genome-wide significant loci associated with alcohol use—providing a robust dataset for testing enrichment of biologically plausible gene sets through partitioned heritability and enrichment analyses.

**Step 1:** We identified and obtained the gene sets. Two prior studies had identified genes in mice associated with binge drinking behaviors that were differentially expressed in rodent models. These are available in the GeneWeaver database provided by K. Marballi ((Marballi, et al., 2016); GSID: GS399061) included 1086 genes and the dataset by Megan K. Mulligan ((Mulligan, et al., 2011); GSID: GS399058) comprised 1393 differentially expressed genes in mice. For clarity in downstream analyses, we refer to GS399061 as the binge_drinking_VTA set and GS399058 as the binge_drinking_Str_PFC_VTA set. Gene sets for both studies were supplied to CONSILIENCE-GWAS.

**Step 2:** We performed partitioned heritability analysis via CONSILIENCE-GWAS using the DPW GWAS summary statistics to assess the contribution of cell-specific functional annotations. Specifically, we incorporated precomputed LD score annotations from publicly available resources, including GTEx, the Roadmap Epigenomics Project, and mouse brain cell-type datasets (Cahoy, et al., 2008), to capture cell-specific regulatory information. These annotations were integrated into the analysis to evaluate whether SNPs were disproportionately enriched in functionally relevant genomic regions associated with the biological processes relevant to the trait.

#### 3.1 Identifying A Priori Gene-Set(s)

To identify gene sets relevant to substance use disorder (SUD), we queried the GeneWeaver database (Baker, et al., 2012); https://www.geneweaver.org), a multi-species functional genomics resource that integrates experimental results across organisms and platforms. GeneWeaver provides access to thousands of curated gene sets from publicly available datasets, particularly those related to neurological and behavioral phenotypes, including alcohol and nicotine exposure. This included transcriptomic data from various species subjected to alcohol or nicotine consumption and exposure. To prepare these data for analysis, we used the biomaRt package (Durinck, et al., 2009) and aligned organism-specific gene identifiers to the latest genome assemblies listed in Table 1. This ensured up-to-date and accurate genomic context for downstream integration. Each curated set of genes was matched to a corresponding GeneWeaver gene set ID and then mapped to the human hg19 (GRCh37) reference genome to enable compatibility with human GWAS summary statistics (Figure 5). This step is essential for harmonizing cross-species gene sets into a unified analysis framework, allowing the investigation of evolutionary conservation of addiction-related pathways.

**Figure 5.**
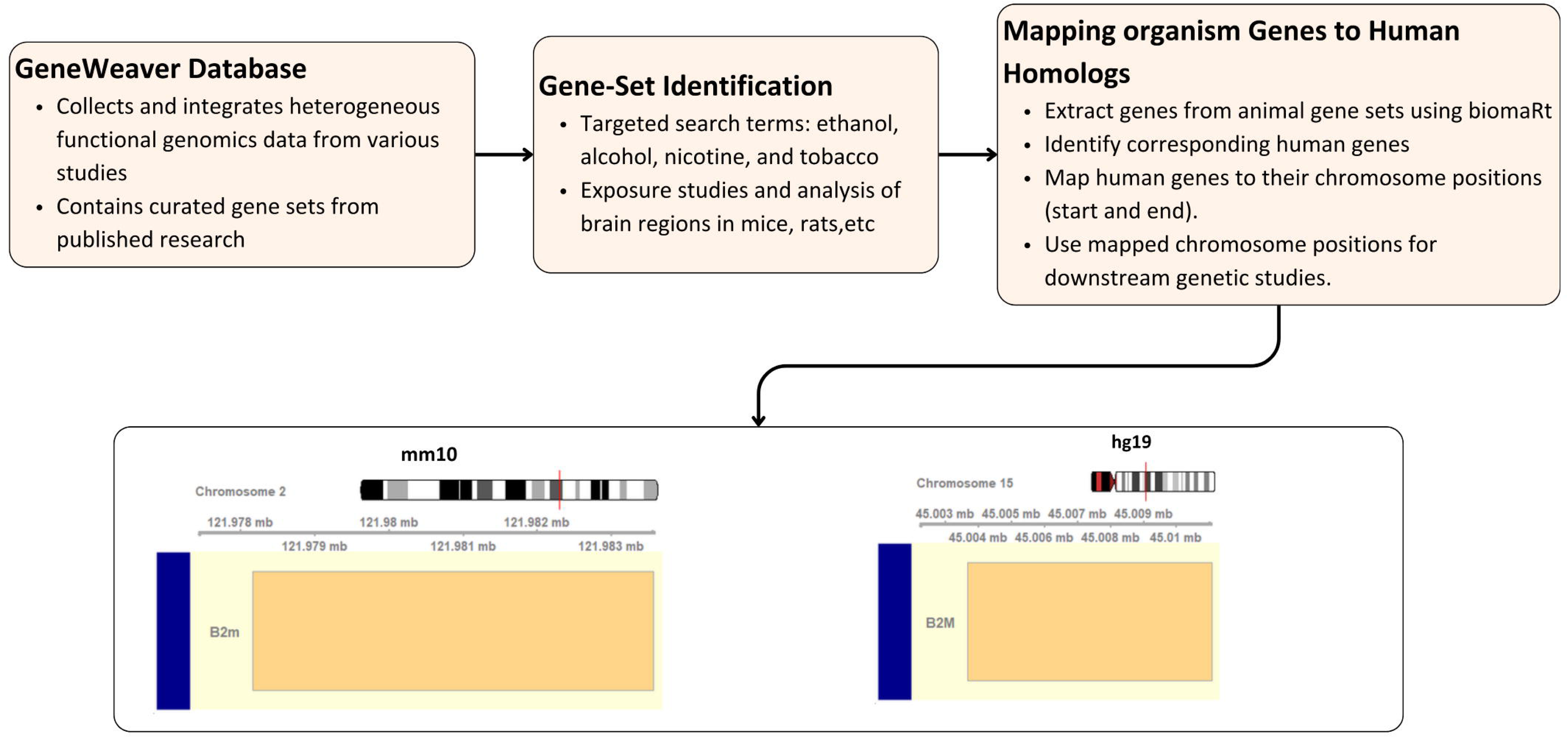
Cross-Species Gene Set Alignment and Mapping for SUD Analysis in CONSILIENCE-GWAS. Mouse sequencing data at the B2m promoter region in the mouse genome (mm10; Chr 2: 121,978,168 - 121,983,563) maps to the B2m gene in human genome (hg19; Chr 15: 44,711,358 - 44,718,851). Transcriptomic data from GeneWeaver are aligned to their respective genome assemblies, mapped to human genome coordinates (hg19) using the biomaRt package, and prepared for downstream enrichment analysis in CONSILIENCE-GWAS.

**Table 1:** Reference Genome Assemblies for Model Organisms and Human.

| **Table 1:**<br>**Reference Genome Assemblies for Model Organisms and Human** |  |
| --- | --- |
| **Species** | **Genome assembly** |
| Mouse (*Mus musculus*) | GRCm39 |
| Rat (*Rattus norvegicus*) | mRatBN7.2 |
| Zebrafish (*Danio rerio*) | GRCz11 |
| Fruit fly (*Drosophila melanogaster*) | BDGP6.46 |
| Dog (*Canis lupus familiaris*) | ROS_Cfam_1.0 |
| (Nematode, N2)<br>(*Caenorhabditis elegans*) | WBcel235 |
| *Saccharomyces cerevisiae*<br>(*Saccharomyces cerevisiae* S288c) | R64-1-1 |
| Macaque (*Macaca mulatta*) | Mmul_10 |
| Human | GRCh37.p13 |
| Note: The genome assemblies listed above are sourced from BioMart reference genome building. |  |

Additionally, we conducted a cell-type analysis using the method described by Finucane (Finucane, et al., 2018). This analysis utilized gene expression data from 205 tissue and cell types, sourced from the GTEx database (Consortium, et al., 2015) and resources provided by the Franke Lab (Pers, et al., 2015), both based on the hg19 genome assembly.

#### 3.2 CONSILIENCE-GWAS Applied Example: Drinks per Week GWAS and Binge Drinking Gene sets

Using the GWASLab Python package (v3.4.46) (He, et al., 2023), we conducted a GWAS for DPW (Figure 6) and identified 41 independent loci in European populations. These loci span genes implicated in synaptic signaling, reward processing, and metabolic function—pathways thought to underline individual differences in alcohol use. To test for functional relevance of gene sets linked to alcohol behavior, we examined whether a binge-drinking-related gene set derived from rodent studies in the GeneWeaver database showed enrichment for DPW heritability. For this purpose, we first used S-LDSC method (Finucane, et al., 2015) to test for enrichment of DPW heritability within this binge-drinking-related gene set as shown in Figure 7a. In this analysis, SNPs were assigned to genes using a ±100 kb window. Our results revealed that the binge-drinking-related gene set was significantly enriched for DPW heritability after correcting for multiple testing (FDR < 0.05), suggesting cross-species relevance of alcohol-associated gene expression patterns. We then extended this analysis using LDSC-SEG to assess cell-type-specific heritability enrichment and found that genes highly expressed in the central nervous system showed the strongest signal, aligning with the neurological underpinnings of alcohol use. Additionally, adrenal/pancreatic, skeletal muscle, and liver cell groups showed significant enrichment after Bonferroni correction (*p* < 0.05), as shown in Figure 7b. These results suggest broader physiological systems may also contribute to individual differences in alcohol intake. Next, we used the LDSC regression approach to see if the heritability of DPW was particularly enriched in the regulatory regions surrounding genes in specific tissues. We observed significant enrichment in promoter-specific epigenetic markers (H3K4me1/me3) in the fetal brain regions, including the germinal matrix and frontal cortex, aided by the multi-tissue chromatin data from ROADMAP and ENCODE as shown in Figure 8.

**Figure 6.**
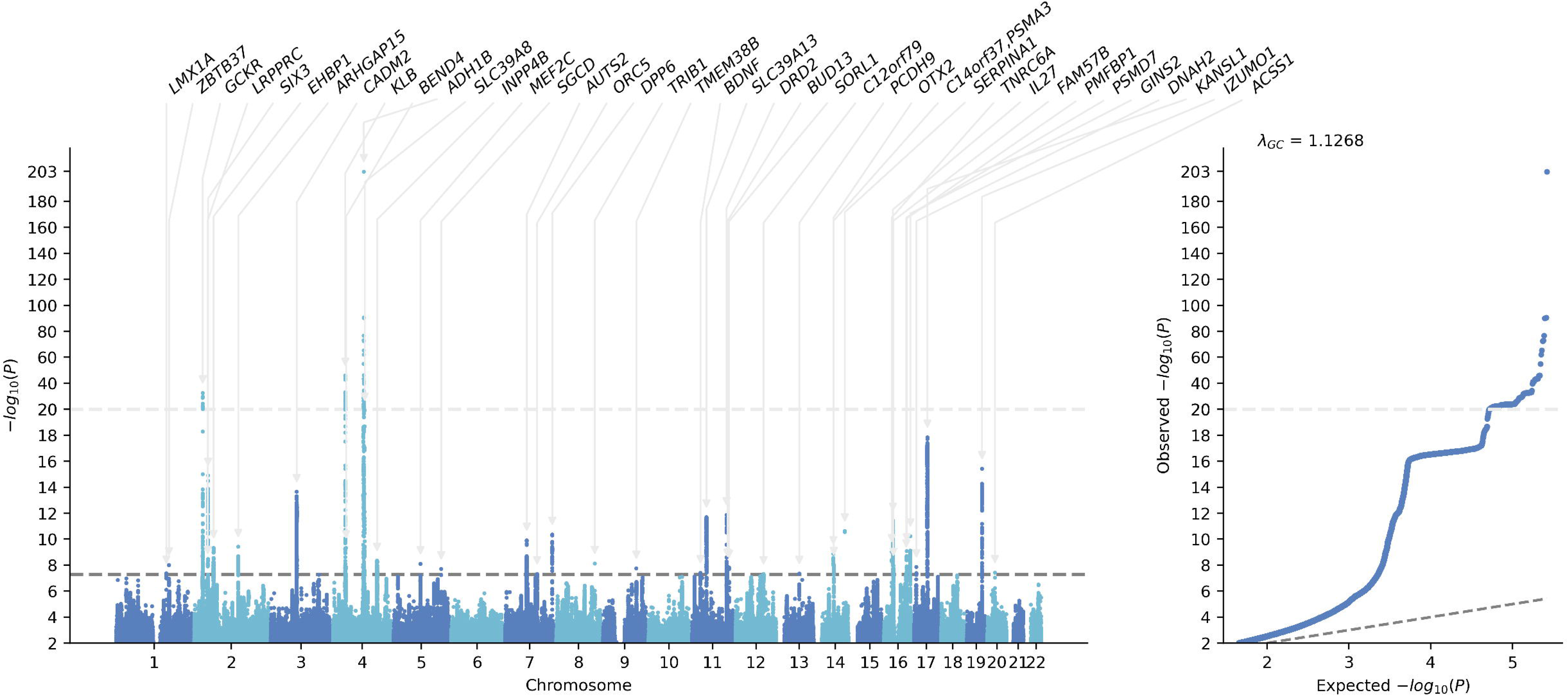
Genome-Wide Association Results for Alcohol Consumption (Drinks per Week) Manhattan plots (left) of Drinks per Week (DPW). QQ plot (right) showing the observed versus expected distribution of p-values under the null hypothesis. Gene names are listed only for the significant variants. The grey line corresponds to p=5×10-8, the standard GWAS significance threshold in individuals of European ancestry.

**Figure 7.**
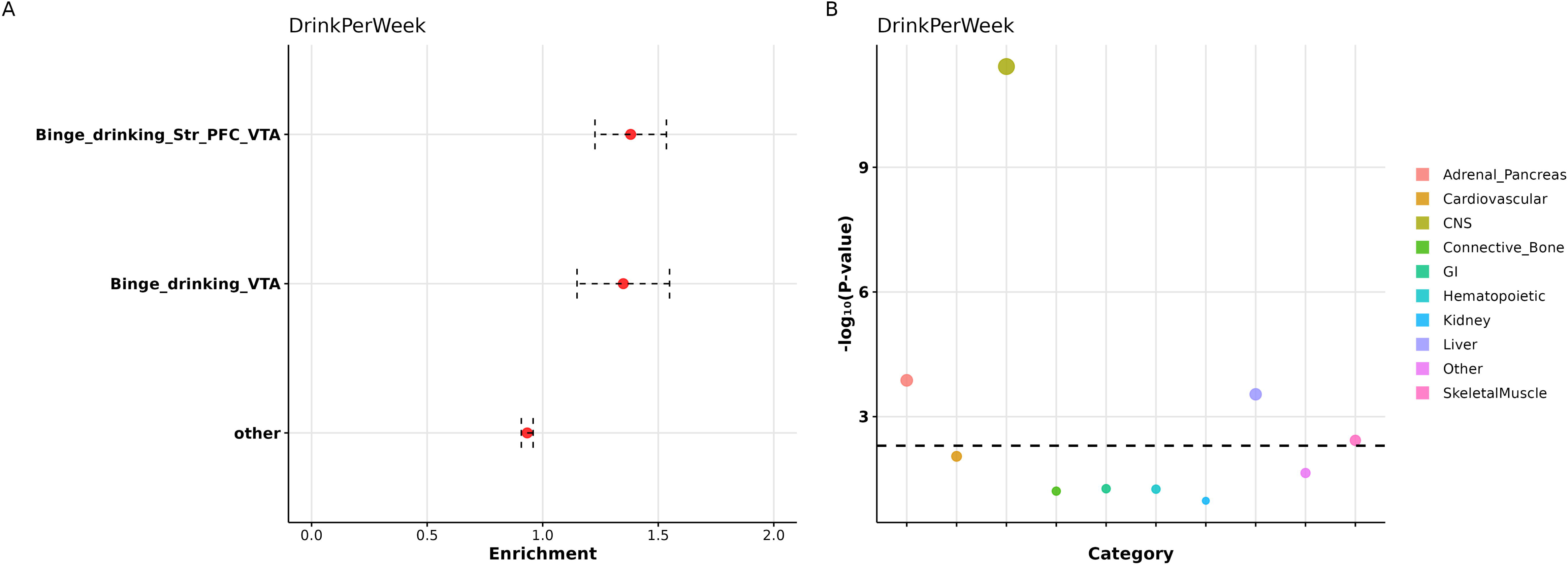
Partitioned Heritability Enrichment of Binge Drinking Gene Sets and CNS Cell Types in Drinks per Week GWAS. The S-LDSC results for DPW studies reveal the enrichment of binge drinking from GeneWeaver mouse gene sets and CNS regions. Each of the 10 enrichment calculations for a particular cell type is performed independently, while controlling for the 53 functional annotation categories in the full baseline model. **A)** Enrichment of binge drinking–related gene sets from GeneWeaver using the DPW GWAS dataset. The horizontal bar displays the enrichment score for binge drinking. **B)** Partitioned heritability enrichment for cell type groups in DPW. Each point represents a tissue/cell type from either the GTEx dataset or the Franke lab dataset. Ten cell types were tested, with corrections for multiple testing. The dashed black lines indicate Bonferroni-corrected significance (P < 0.05/10).

**Figure 8.**
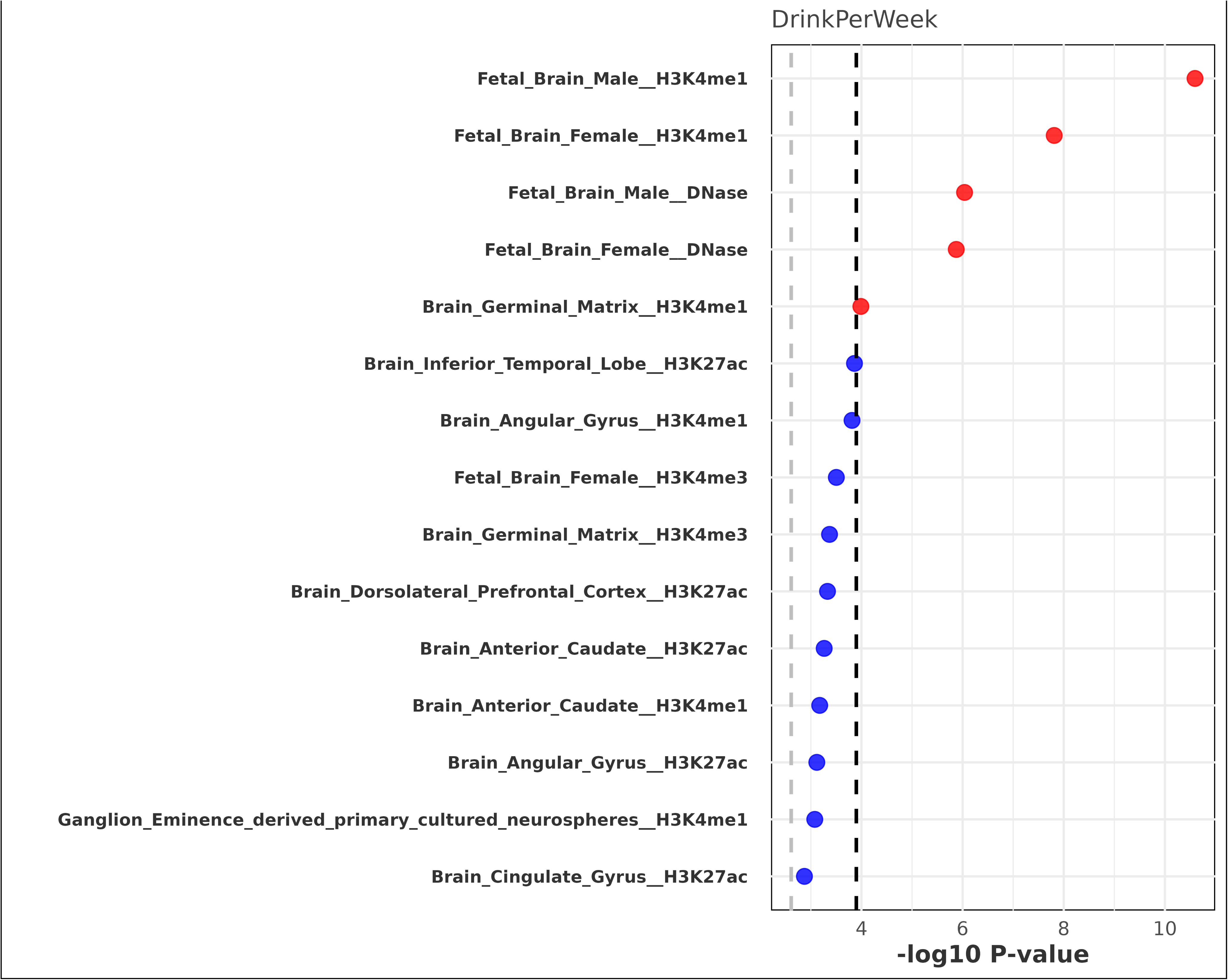
LDSC analysis using tissue specific data. Epigenetic partitioning heritability enrichment for DPW GWAS using Roadmap data. LDSC analysis showed significant enrichment of promoter-specific markers (H3K4me1/me3) in the fetal and adult brain for the SNPs identified in DPW. X-axis represents the annotations, and Y-axis represents the −log 10LP value for enrichment calculated using partition heritability method as implemented in LDSC. The dotted black line represents the threshold of multiple test correction according to Bonferroni. The gray dashed line is the cutoff for FDRL<L0.05.

**Figure 9.**
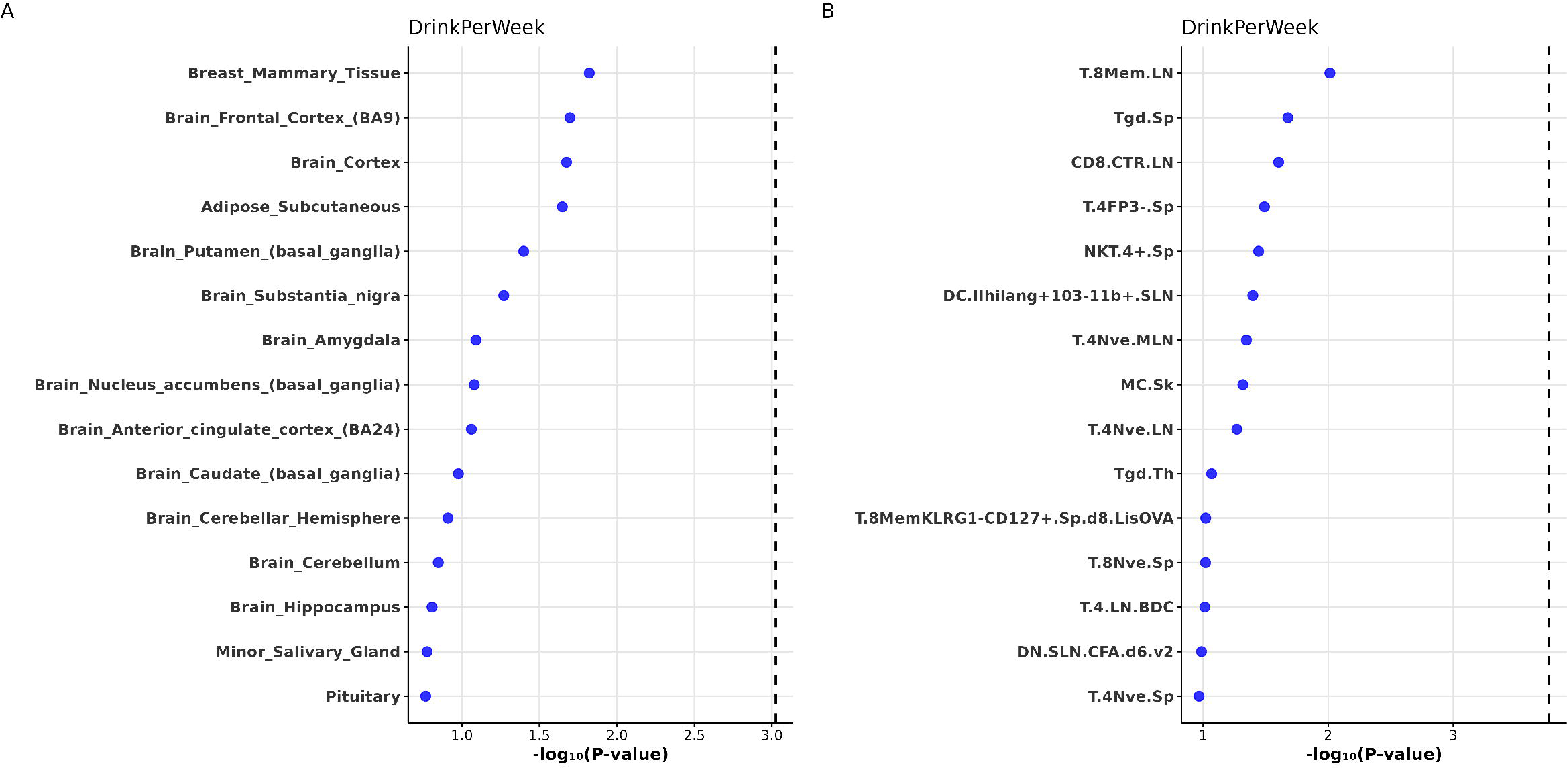
Partitioned Heritability Enrichment of Drinks per Week (DPW) Using Human and Mouse Gene Expression Data. **A)** Tissue or cell type partitioning heritability enrichment for DPW GWAS using GTEx gene expression data. **B)** Mouse immune cell type partitioning heritability enrichment DPW using ImmGen gene expression data.

#### 3.3 Joint Model of Enrichment Analysis of Binge Drinking-Associated Gene sets

We applied S-LDSC to assess the enrichment of SNP-based heritability for binge drinking within gene sets derived from rodent models of alcohol exposure, with a particular focus on transcriptional activity in brain regions involved in reward processing, including the prefrontal cortex (PFC), striatum, and ventral tegmental area (VTA). In the joint model (Table 2), which simultaneously included all candidate gene sets while adjusting for baseline LD annotations, we observed significant enrichment for the *Binge_drinking_Str_PFC_VTA_UniqueGenes* set. This set, comprising genes uniquely expressed in the striatum-PFC-VTA and excluding those overlapping with other sets, explained 14.5% of SNP heritability despite accounting for only 10.3% of analyzed SNPs (enrichment = 1.41) indicating strong region-specific genetic contribution. The *Binge_drinking_VTA_UniqueGenes* set, consisting of VTA-specific genes demonstrated significant enrichment, albeit to a lesser extent (enrichment = 1.35). In contrast, the intersecting gene sets (*Binge_drinking_Str_PFC_VTA_and_Binge_drinking_VTA_IntersectionGenes*) showed modest enrichment (enrichment = 1.61) that did not survive multiple testing correction, possibly reflecting limited statistical power or divergent regulatory roles across regions. SNPs outside these defined sets (other_Variants) showed no evidence of enrichment (i.e., enrichment ≈ 1.00).

**Table 2.**
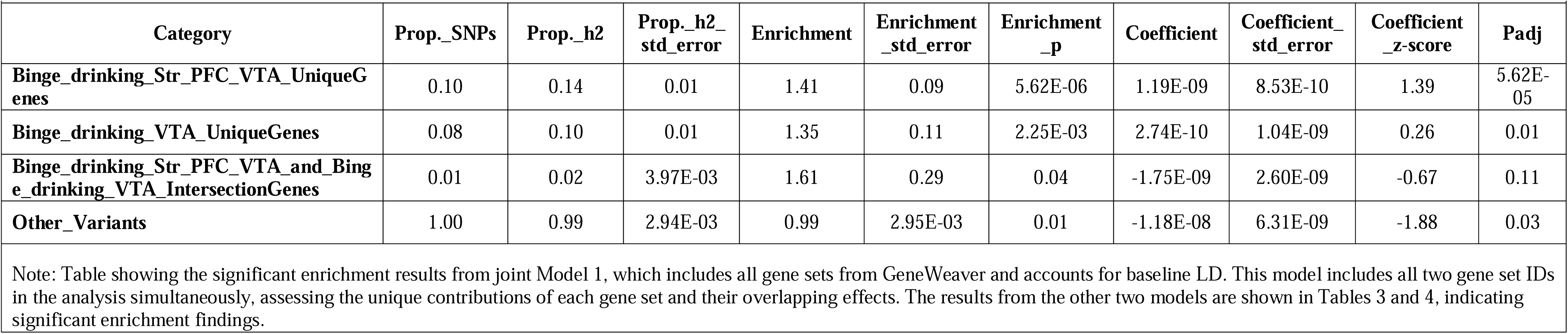
Partitioned Heritability of Binge Drinking in Joint Gene Set Model.

To further dissect the contribution of these gene sets, we examined two additional models: a union model and a region-specific model. In the union model (Table 3), which collapsed all binge drinking-related genes into a single annotation (*All_UnionGenes*), we observed robust enrichment (enrichment = 1.32), with 23.0% of SNP heritability explained by just 17.5% of SNPs. This finding supports the cumulative relevance of these cross-species, functionally characterized gene sets to binge drinking phenotypes. In the region-specific model (Table 4), we evaluated two gene sets obtained directly from the GeneWeaver database: *Binge_drinking_Str_PFC_VTA* and *Binge_drinking_VTA* both derived from rodent brain transcriptional profiles following binge alcohol exposure. The *Binge_drinking_Str_PFC_VTA* set again showed significant enrichment (enrichment = 1.38), accounting for 15.7% of heritability from 11.4% of SNPs. The *Binge_drinking_VTA* set showed nominal enrichment (enrichment = 1.35). While these two sets were analyzed separately, they may partially overlap due to shared transcriptional responses across brain regions. This nuance should be considered when interpreting the specificity of enrichment estimates.

**Table 3.** Enrichment of Binge Drinking SNP Heritability in Union of All Rodent Gene Sets.

| Category | Prop._SNPs | Prop._h2 | Prop._h2_std_error | Enrichment | Enrichment_std_error | Enrichment_p | Coefficient | Coefficient_std_error | Coefficient_z-score | Padj |
| --- | --- | --- | --- | --- | --- | --- | --- | --- | --- | --- |
| All_UnionGenes | 0.17 | 0.23 | 0.01 | 1.32 | 0.06 | 9.86E-07 | 1.56E-05 | 1.17E-05 | 1.34 | 1.07E-05 |
| other_Variants | 0.83 | 0.77 | 0.01 | 0.93 | 0.01 | 9.86E-07 | 1.56E-05 | 1.17E-05 | 1.34 | 1.07E-05 |

Note: This table shows results from a stratified LD score regression model where all candidate genes identified from rodent binge drinking studies were combined into a single union annotation

**Table 4.** Region-Specific Enrichment of Binge Drinking SNP Heritability Using Gene Sets from Striatum-PFC-VTA and VTA (PMID IDs: 21223303, 26482798)

| Category | Prop._SNPs | Prop._h2 | Prop._h2_std_error | Enrichment | Enrichment_std_error | Enrichment_p | Coefficient | Coefficient_std_error | Coefficient_z-score | Padj |
| --- | --- | --- | --- | --- | --- | --- | --- | --- | --- | --- |
| **Binge_drinking_Str_PFC_VTA** | 0.11 | 0.16 | 0.01 | 1.38 | 0.08 | 3.30E-06 | 2.79E-05 | 1.32E-05 | 2.12 | 2.94E-05 |
| **other_Variants** | 0.89 | 0.84 | 0.01 | 0.95 | 0.01 | 3.30E-06 | 2.79E-05 | 1.32E-05 | 2.12 | 2.94E-05 |
| **Binge_drinking_VTA** | 0.09 | 0.12 | 0.01 | 1.35 | 0.10 | 1.06E-03 | -1.82E-05 | 2.12E-05 | -0.86 | 5.78E-03 |
| **other_Variants** | 0.91 | 0.88 | 0.01 | 0.97 | 0.01 | 1.06E-03 | -1.82E-05 | 2.12E-05 | -0.86 | 5.78E-03 |

Note: This table reports results from region-specific S-LDSC analyses using two gene sets derived directly from the GeneWeaver database. One set includes genes from the striatum, PFC, and VTA (ID: GS399058), and the other includes VTA-specific genes (ID: GS399061).

Overall, in the joint model, both the striatum–PFC–VTA and VTA-specific unique gene sets reached significance after FDR correction (FDR < 0.05). In the union and region-specific models, the All_UnionGenes set, as well as the striatum–PFC–VTA and VTA-specific sets, showed significant enrichment after FDR correction. Together, these findings highlight the relevance of transcriptional regulation in the PFC and VTA to the genetic architecture of binge drinking and demonstrate the power of integrating rodent-derived functional genomic data with human GWAS to uncover biologically meaningful, tissue-specific contributions to complex behavioral traits.

#### 3.4 Gene Set-Based PGS Application

We also employed PRS-CS -auto to generate gene set-specific PGS by calculating posterior SNP effect sizes (adjusted beta weights) based on GWAS summary statistics. Gene sets were derived from the GeneWeaver database and reflected transcriptional responses to binge alcohol exposure in rodent brain regions implicated in reward processing. Specifically, we constructed PGS for (1) genes expressed in the striatum, prefrontal cortex (PFC), and ventral tegmental area (VTA) associated with binge drinking, (2) VTA-specific genes related to binge alcohol exposure, (3) a union gene set combining all binge drinking-relevant genes across regions (PGS_All_UnionGenes), and (4) a set of background SNPs not overlapping with the defined gene sets (PGS_other). This approach enabled us to partition polygenic signal across biologically informed annotations and quantify the relative contribution of each functionally derived gene set to the genetic architecture of the trait. The use of PRS-CS allows for shrinkage-based correction of GWAS effect sizes, thus enhancing the interpretability of gene set-level polygenic risk profiles.

We created polygenic scores (PGSs) for DPW in the European ancestry subset of the QIMR Berghofer h (QIMR; N=24,732). Participants were genotyped using commercial arrays that can be broken into 5 batches (1^st^ batch: Illumina 610K, 660K, 2^nd^ batch: Illumina 370K, 317K, 3^rd^ batch: CoreExome, PsychArray, PsychChip, 4^th^ batch: OmniExpress_2.5M, and 5^th^ batch: GSA). Details of QC and imputation are available in the Supplemental Materials. After estimating posterior SNP effect sizes via PRS-CS, PGSs were computed using PLINK 1.9 --score function for the individuals of European ancestry only. The association between the DPW PGS and typical alcohol consumption (measured as the number of drinks per day) was evaluated across four QIMR cohorts: the 25UP Study (Mitchell, et al., 2019) (N = 1,952), Genetic Epidemiology of Pathological Gambling (Slutske, et al., 2009) (GA, N = 677), Nicotine Project (Saccone, et al., 2007) (NC, N = 1,080), and the Semi-Structured Assessment for the Genetics of Alcoholism Phase 1 (Heath, et al., 2011) (SS1, N =4254). Linear regression models in R were used, adjusting for age, sex, and the first four within-ancestry principal components.

##### Polygenic Effects on Alcohol Consumption

Table 5 summarizes the results of the regression models examining the effects of each polygenic score (PGS) on alcohol consumption (log drinks per day). In the independent models, where each PGS is tested individually, all PGSs were significantly associated with increased alcohol consumption. The largest individual gene set effect was observed for PGS_other category (β = 0.057, 95% CI = [0.047, 0.067], p < 0.001).

**Table 5.**
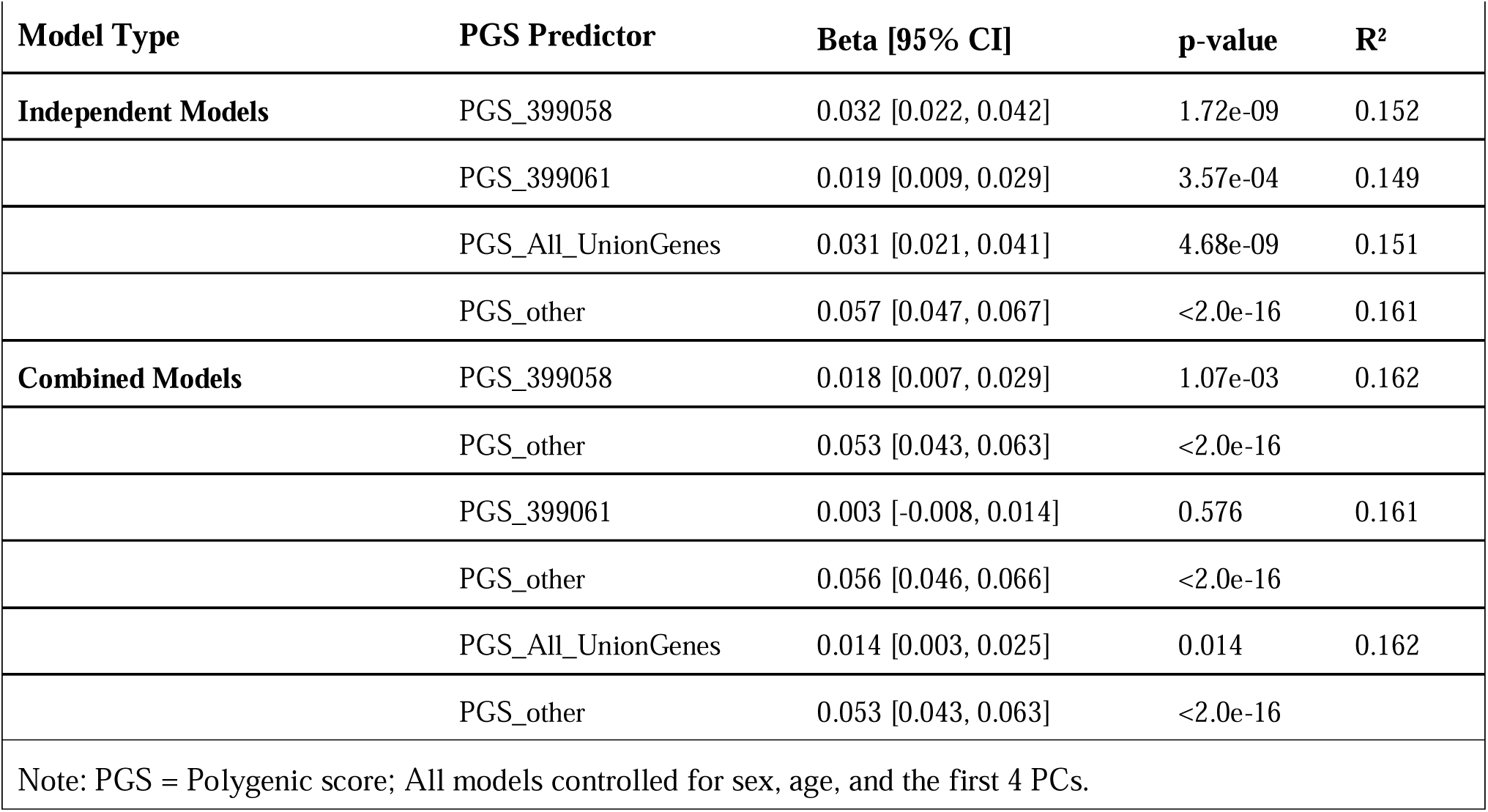
Association between PGS and Alcohol Consumption (log drinks per day + 1)

When the effects of the PGS_other were incorporated into the combined models, where all PGSs were tested simultaneously, we observed a key distinction in the unique contributions of the other scores. The PGS_All_UnionGenes and the PGS 399058 continued to show significant, albeit attenuated, positive associations with alcohol consumption (PGS All Union Genes: β = 0.014, 95% CI = [0.002, 0.025], p = 0.014; PGS 399058: β = 0.018, 95% CI = [0.007, 0.029], p = 0.0011), indicating they capture unique genetic influences independent of the PGS Other. In contrast, the association for PGS 399061 was fully attenuated and became non-significant in the combined model (β = 0.003, 95% CI = [-0.008, 0.014], p = 0.576), suggesting its effect is not unique from the PGS Other.

No evidence of multicollinearity was detected in the combined models (VIF < 2 for all predictors).

## 4. Discussion

We present CONSILIENCE-GWAS, a comprehensive, web-based platform that advances the integration of functional genomic data from animal models with human GWAS findings. The tool automates critical steps in heritability enrichment analysis—including stratified LD Score Regression, polygenic score calculation, and joint modeling—within a user-friendly interface accessible to researchers without computational expertise. By allowing seamless upload of GWAS summary statistics and direct entry of GeneWeaver cross-species gene set IDs (without preprocessing), CONSILIENCE-GWAS removes many of the technical barriers historically associated with cross-species analysis. The platform supports visualization of cell- and tissue-specific heritability enrichment patterns (Finucane, et al., 2018) enabling users to interpret the genetic architecture of complex traits through high-resolution enrichment plots. These capabilities ensure standardized, reproducible workflows and improve accessibility for both novice and experienced users.

CONSILIENCE-GWAS represents a transformative advancement in translational genomics by enabling researchers to evaluate evolutionarily conserved biological mechanisms underlying complex traits. In a demonstration analysis using GWAS of alcohol consumption and rodent binge-drinking gene sets, the platform revealed significant enrichment in striatal-prefrontal-ventral tegmental regions (enrichment = 1.41, *p* = 5.62×10LL), supporting its utility in uncovering biologically meaningful signals. Overall, by combining enrichment analysis, polygenic scoring, and visualization in a centralized platform, CONSILIENCE-GWAS helps connect findings from animal and human genetics, making it easier to discover treatment targets for conditions like substance use disorders, where many genes work similarly across species.

CONSILIENCE-GWAS adds to a growing landscape of integrative tools in the field of psychiatric genetics. Existing platforms such as FUMA provide powerful functionality for mapping GWAS loci to candidate genes using positional, eQTL, and chromatin interaction mapping, along with gene-based, tissue-specific, and pathway-level annotations that help prioritize biological mechanisms underlying genetic associations (Watanabe, et al., 2017). Another widely used resource is the Complex-Traits Genetics Virtual Lab (CTG-VL) that integrates a comprehensive suite of tools for the downstream analysis of GWAS summary statistics, offering functions for estimating heritability and genetic correlations, performing gene-based and interactively visualizing results (Cuéllar-Partida, et al., 2019). Importantly, CTG-VL also aggregates pre-computed results for more than 1,500 traits, enabling researchers to explore pleiotropy and generate biological hypotheses efficiently. However, these existing resources are primarily focused on analyses within the human genome. CONSILIENCE-GWAS distinguishes itself by providing the first web-based platform specifically designed for hypothesis-driven, cross-species functional enrichment analysis. Unlike other tools that permit custom gene set uploads, CONSILIENCE-GWAS is uniquely integrated with the GeneWeaver database (Baker, et al., 2012), offering direct access to thousands of curated gene sets from model organisms without requiring manual preprocessing. This seamless integration allows researchers to test, for the first time in an automated workflow, whether genes implicated in animal models of a trait (e.g., binge drinking in mice) explain a significant portion of the heritability of the corresponding human trait (e.g., alcohol consumption). Furthermore, CONSILIENCE-GWAS evaluates heritability proportions for specific functional categories and identifies tissue- and cell-type specificity using stratified LD score regression (S-LDSC) (Finucane, et al., 2018). By bringing together stratified LDSC, polygenic scoring with PRS-CS (Ge, et al., 2019) and cross-species gene set analysis in one easy-to-use platform, CONSILIENCE-GWAS bridges the gap between animal studies and human genetics. It provides a unique resource that helps researchers translate discoveries from model organisms into clearer insights about the genetic basis of human complex traits.

As GWAS continues to increase in power, we anticipate greater need for integration approaches to interpret variant associations using experimentally relevant datasets. In the future, we plan to enhance this resource by expanding the LD reference panels to include populations that are underrepresented in GWAS. Although cross-ancestry GWASs do minimize the degree of bias from European-centered studies, there remain concerns about the relative power of GWAS in populations of African, East and South Asian, and Latin-American groups. Current efforts by the International Society of Psychiatric Genetics (LázaroLMuñoz, et al., 2019) and the Polygenic Risk Methods in Diverse Populations (PRIMED) Consortium (Kullo, et al., 2024) are expected to reduce this disparity.

### Conclusions

CONSILIENCE-GWAS offers a powerful and user-friendly platform for integrating gene sets from model organisms with human GWAS data to explore cross-species enrichment in complex traits. By combining GeneWeaver gene sets with stratified LD score regression and polygenic scoring, the tool can detect significant enrichment of model organism findings in human studies. Its automated workflow and web interface make these analyses accessible to researchers without advanced computational skills. Although currently optimized for European ancestry, future updates will expand its utility to more diverse populations and finer-resolution data, advancing our ability to translate insights from animal models to human health.

## Supporting information

main manuscript

## Data Availability

All data used in this study are publicly available GWAS summary statistics
and functional genomic datasets. The CONSILIENCE-GWAS web platform is
available at https://consilience.bgalab.emory.edu/

## 5. Availability

Consilience-GWAS is accessible at: https://consilience.bgalab.emory.edu

## Acknowledgements

We would like to thank the extant researchers in the field who have contributed to the field of addiction science and openly share their data via platforms sponsored by the National Institutes of Health, such as GeneWeaver, and the Mouse Phenome Database, or through their own efforts, without which none of this work would be possible.

The Genetic Epidemiology of Pathological Gambling (GA) study was established by Wendy S. Slutske and Nicholas G. Martin and supported by National Institutes of Health Grants (MH66206; AA013320; AA013321; AA013326; AA014041; AA011998; AA017688; DA012854). A portion of the genotyping on which this study was based (Illumina HumanCNV370 scans on 624 individuals) as carried out at the Center for Inherited Disease Research, Baltimore, through an access award to our late colleague Dr Richard Todd (Psychiatry,Washington University School of Medicine, St Louis). We appreciate the continued participation of the Australian Twin Registry twins. We also thank Bronwyn Morris, Megan Fergusson, David Smyth, Olivia Zheng, and Harry Beeby for data collection and data management.

The Nicotine Project (NC) was an international collaborative study established by Nicholas G. Martin, Andrew C. Heath and Pamela AF Madden. The NC study was supported by National Institutes of Health grants DA12854, AA07728, AA07580, AA13321, AA13320, and DA019951. Funding was also provided by the Australian National Health and Medical Research Council (241944, 339462, 389927, 389875, 389891, 389892, 389938, 442915, 442981, 496739, 552485, 552498). A portion of the genotyping on which this study was based was carried out at the Center for Inherited Disease Research, Baltimore (CIDR) (Illumina 370K scans on 4300 individuals), through an access award to our late colleague Dr. Richard Todd. We warmly thank the participating twin pairs and their family members for their contribution. We also thank Bronwyn Morris, Megan Fergusson, David Smyth, Olivia Zheng, Harry Beeby, Anjali Henders, Dixie Statham, Richard Parker, Soad Hancock, Judith Moir, Sally Rodda, Pieta-Maree Shertock, Heather Park, Jill Wood, Pam Barton, Fran Husband, and Adele Somerville for data collection and data management.

The 25UP study (TU) was funded by an Australian National Health and Medical Research Council Project Grant (No. APP1069141) to Ian B. Hickie and Nicholas G. Martin. We thank the twins and their families for their participation in our research and David Smyth and Scott Gordon for IT support.

The Semi-Structured Assessment for the Genetics of Alcoholism Phase 1 (SS1) study was established by Nicholas G. Martin, Andrew C. Heath and Pamela AF Madden. The SS1 study was supported by National Institutes of Health Grants AA07535, AA07728, AA13320, AA13321, AA14041, AA11998, AA17688, DA012854, and DA019951; by Grants from the Australian National Health and Medical Research Council (241944, 339462, 389927, 389875, 389891, 389892, 389938, 442915, 442981, 496739, 552485, and 552498); by Grants from the Australian Research Council (A7960034, A79906588, A79801419, DP0770096, DP0212016, and DP0343921); and by the 5th Framework Programme (FP-5) GenomEUtwin Project (QLG2-CT-2002-01254). Genome-wide Association study genotyping at Center for Inherited Disease Research was supported by a Grant to the late Richard Todd, M.D., Ph.D., former Principal Investigator of Grant AA13320 and a key contributor to research described in this article. We acknowledge the contributions of project investigator Alexandre Todorov, Ph.D., at Washington University. We also thank Dixie Statham, Ann Eldridge, Marlene Grace, Kerrie McAloney (sample collection); Lisa Bowdler and Steven Crooks (DNA processing); and David Smyth, Harry Beeby, and Daniel Park (Information Technology support) at QIMR Berghofer, Brisbane Australia. Last, but not least, we thank the twins and their families for their participation.

## Data availability

All of the GWAS summary data used in the current example are publicly available. Access can be gained from the journal website for each paper or the Database for Genotypes and Phenotypes. Geneweaver gene sets are accessible at Geneweaver.org. Further information on the CONSILIENCE-GWAS pipeline is also available on the CONSILIENCE-GWAS website using the Overview and Tutorial pages.

## References

A haplotype map of the human genome. Nature 2005;437(7063):1299–1320.

Agrawal, A., et al. The genetics of addiction—a translational perspective. Translational Psychiatry 2012;2(7):e140–e140.

Anderson, C.A., et al. Data quality control in genetic case-control association studies. Nat Protoc 2010;5(9):1564–1573.

Baker, E., et al. GeneWeaver: data driven alignment of cross-species genomics in biology and disease. Nucleic acids research 2016;44(D1):D555–D559.

Baker, E.J., et al. GeneWeaver: a web-based system for integrative functional genomics. Nucleic Acids Res 2012;40(Database issue):D1067–1076.

Benca-Bachman, C.E., et al. Polygenic influences on the behavioral effects of alcohol withdrawal in a mixed-ancestry population from the collaborative study on the genetics of alcoholism (COGA). Mol Cell Neurosci 2023;125:103851.

Bogue, M.A., et al. Mouse Phenome Database: towards a more FAIR-compliant and TRUST-worthy data repository and tool suite for phenotypes and genotypes. Nucleic acids research 2023;51(D1):D1067–D1074.

Bogue, M.A., et al. Mouse phenome database: curated data repository with interactive multi-population and multi-trait analyses. Mammalian Genome 2023;34(4):509–519.

Brasher, M.S., et al. Testing associations between human anxiety and genes previously implicated by mouse anxiety models. Genes Brain Behav 2023;22(6):e12851.

Bulik-Sullivan, B.K., et al. LD Score regression distinguishes confounding from polygenicity in genome-wide association studies. Nature Genetics 2015;47(3):291–295.

Cahoy, J.D., et al. A transcriptome database for astrocytes, neurons, and oligodendrocytes: a new resource for understanding brain development and function. Journal of Neuroscience 2008;28(1):264–278.

Consortium, E.P. An integrated encyclopedia of DNA elements in the human genome. Nature 2012;489(7414):57.

Consortium, G., et al. The Genotype-Tissue Expression (GTEx) pilot analysis: multitissue gene regulation in humans. Science 2015;348(6235):648–660.

Consortium, G.P. A global reference for human genetic variation. Nature 2015;526(7571):68.

Cuéllar-Partida, G., et al. Complex-Traits Genetics Virtual Lab: A community-driven web platform for post-GWAS analyses. bioRxiv 2019:518027.

Durinck, S., et al. BioMart and Bioconductor: a powerful link between biological databases and microarray data analysis. Bioinformatics 2005;21(16):3439–3440.

Durinck, S., et al. Mapping identifiers for the integration of genomic datasets with the R/Bioconductor package biomaRt. Nature Protocols 2009;4(8):1184–1191.

Finucane, H.K., et al. Partitioning heritability by functional annotation using genome-wide association summary statistics. Nature Genetics 2015;47(11):1228–1235.

Finucane, H.K., et al. Partitioning heritability by functional annotation using genome-wide association summary statistics. Nat Genet 2015;47(11):1228–1235.

Finucane, H.K., et al. Heritability enrichment of specifically expressed genes identifies disease-relevant tissues and cell types. Nature Genetics 2018;50(4):621–629.

Ge, T., et al. Polygenic prediction via Bayesian regression and continuous shrinkage priors. Nature communications 2019;10(1):1776.

He, Y., et al. GWASLab: a Python package for processing and visualizing GWAS summary statistics. 2023.

Heath, A.C., et al. A quantitative-trait genome-wide association study of alcoholism risk in the community: findings and implications. Biological psychiatry 2011;70(6):513–518.

Huggett, S.B., et al. Genes identified in rodent studies of alcohol intake are enriched for heritability of human substance use. Alcohol Clin Exp Res 2021;45(12):2485–2494.

Huggett, S.B., et al. Bulk and Single-cell Transcriptomic Brain Data Identify Overlapping Processes and Cell-types with Human AUD and Mammalian Models of Alcohol Use. bioRxiv 2024.

Kullo, I.J., et al. The PRIMED Consortium: Reducing disparities in polygenic risk assessment. The American Journal of Human Genetics 2024;111(12):2594–2606.

Kundaje, A., et al. Roadmap Epigenomics Consortium: integrative analysis of 111 reference human epigenomes. Nature;518:317.

LázaroLJMuñoz, G., et al. International society of psychiatric genetics ethics committee: issues facing us. American Journal of Medical Genetics Part B: Neuropsychiatric Genetics 2019;180(8):543–554.

Li, A., et al. Benchmarking methods integrating GWAS and single-cell transcriptomic data for mapping trait-cell type associations. medRxiv 2025:2025.2005.2024.25328275.

Liu, M., et al. Association studies of up to 1.2 million individuals yield new insights into the genetic etiology of tobacco and alcohol use. Nature Genetics 2019;51(2):237–244.

Mailman, M.D., et al. The NCBI dbGaP database of genotypes and phenotypes. Nature genetics 2007;39(10):1181–1186.

Marballi, K., et al. Alcohol consumption induces global gene expression changes in VTA dopaminergic neurons. Genes Brain Behav 2016;15(3):318–326.

Mitchell, B.L., et al. Twenty-five and up (25Up) study: a new wave of the Brisbane Longitudinal Twin Study. Twin Research and Human Genetics 2019;22(3):154–163.

Mulligan, M.K., et al. Molecular profiles of drinking alcohol to intoxication in C57BL/6J mice. Alcohol Clin Exp Res 2011;35(4):659–670.

Palmer, R.H., et al. Integration of evidence across human and model organism studies: A meeting report. Genes, Brain and Behavior 2021;20(6):e12738.

Papatheodorou, I., et al. Expression Atlas update: from tissues to single cells. Nucleic acids research 2020;48(D1):D77–D83.

Pers, T.H., et al. Biological interpretation of genome-wide association studies using predicted gene functions. Nature communications 2015;6(1):5890.

Pers, T.H., et al. Biological interpretation of genome-wide association studies using predicted gene functions. Nature Communications 2015;6(1):5890.

Privé, F., Arbel, J. and Vilhjálmsson, B.J. LDpred2: better, faster, stronger. Bioinformatics 2020;36(22-23):5424–5431.

Purcell, S., et al. PLINK: a tool set for whole-genome association and population-based linkage analyses. The American journal of human genetics 2007;81(3):559–575.

Ramasamy, A., et al. Genetic variability in the regulation of gene expression in ten regions of the human brain. Nature neuroscience 2014;17(10):1418–1428.

Saccone, S.F., et al. Genetic linkage to chromosome 22q12 for a heavy-smoking quantitative trait in two independent samples. The American Journal of Human Genetics 2007;80(5):856–866.

Slutske, W.S., et al. The Australian Twin Study of Gambling (OZ-GAM): rationale, sample description, predictors of participation, and a first look at sources of individual differences in gambling involvement. Twin Research and Human Genetics 2009;12(1):63–78.

Tryka, K.A., et al. NCBI’s Database of Genotypes and Phenotypes: dbGaP. Nucleic acids research 2014;42(D1):D975–D979.

Watanabe, K., et al. Functional mapping and annotation of genetic associations with FUMA. Nature communications 2017;8(1):1826.

Yang, J., et al. GCTA: a tool for genome-wide complex trait analysis. The American Journal of Human Genetics 2011;88(1):76–82.

