## Supplementary material for "CONSILIENCE-GWAS: A Web Resource for Parsed Heritability & Polygenic Score Analysis of Human GWAS Using Heterogeneous Functional Genomics Data": main manuscript

**Supplemental Materials**

***QIMR Sample Description, Genetic QC, & Imputation***

Genotype data were managed and cleaned using PLINK (Purcell, et al., 2007) prior to imputation. To ensure consistency across arrays, variant IDs and position information were aligned to a common reference genome build, Genome Reference Consortium Human build 37 (GRCh37) (Church, et al., 2011). Prior to imputation, we performed quality control by removing SNPs with a call rate of <95%, low Minor Allele Frequency (MAF <1%) on each batch.

Genotype imputation was performed on the Michigan Imputation Server (Das, et al., 2016) https://imputationserver.sph.umich.edu using ShapeIT [v2.r790] (Howie, et al., 2012) for phasing and Minimac3 (Das, et al., 2016). Haplotype Research Consortium (HRC.r1-1) ‘EUR-population’ was used as the reference panel (McCarthy, et al., 2016). The average imputation quality score by minor allele frequency for each platform is shown in Figure 1. The imputation quality was very similar across all five batches. SNPs with an imputation accuracy of <0.5 were excluded. Each dataset was imputed individually based on site and platform and then merged based on the set of intersecting markers to get only high-quality markers from all the imputed data (Winkler, et al., 2014). Analyses also excluded variants with a call rate <90%, (2) p < 1 × 10−4 in Hardy–Weinberg equilibrium tests, and (3) minor allele frequency (MAF <5%) across the whole sample.

The breakdown of 5 batches is (1) Illumina 610K_660K (n = 4007, SNP = 503022); (2) Illumina 370K_317K (n = 3400 , SNP = 283595 ); (3) CoreExome_PsychArray_PsychChip (n = 3243 , SNP = 249693 ); (4) OmniExpress_2.5M (n = 642 , SNP = 606262); (5) GSA (n = 12452,SNP = 432234 )

**Figure 1. Imputation quality across genotyping arrays from QIMR data**

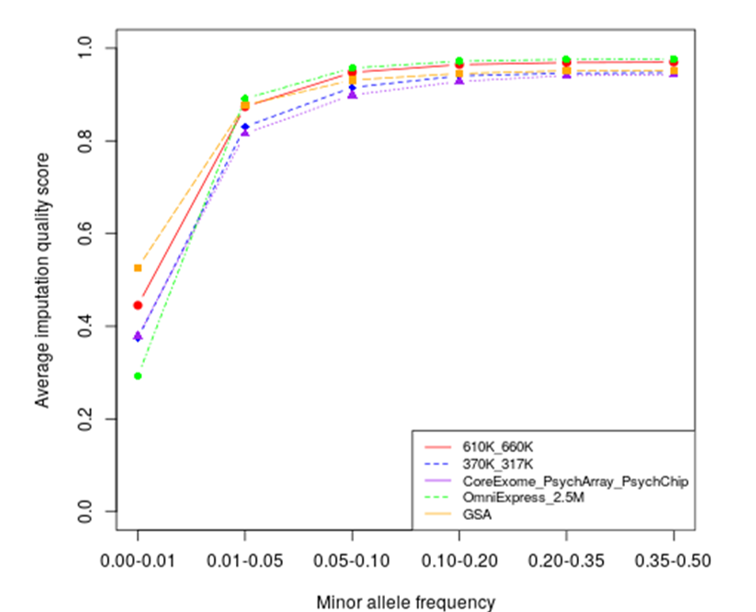

**References**

Church, D.M.*, et al.* Modernizing reference genome assemblies. *PLoS biology* 2011;9(7):e1001091.

Das, S.*, et al.* Next-generation genotype imputation service and methods. *Nature genetics* 2016;48(10):1284–1287.

Howie, B.*, et al.* Fast and accurate genotype imputation in genome-wide association studies through pre-phasing. *Nature genetics* 2012;44(8):955–959.

McCarthy, S.*, et al.* C. the Haplotype Reference. *A reference panel of 64,976 haplotypes for genotype imputation* 2016:1279–1283.

Purcell, S.*, et al.* PLINK: a tool set for whole-genome association and population-based linkage analyses. *The American journal of human genetics* 2007;81(3):559–575.

Winkler, T.W.*, et al.* Quality control and conduct of genome-wide association meta-analyses. *Nature protocols* 2014;9(5):1192–1212.
